# Diagnostic accuracy of two commercial SARS-CoV-2 Antigen-detecting rapid tests at the point of care in community-based testing centers

**DOI:** 10.1101/2020.11.20.20235341

**Authors:** Alice Berger, Marie Therese Ngo Nsoga, Francisco Javier Perez-Rodriguez, Yasmine Abi Aad, Pascale Sattonnet-Roche, Angèle Gayet-Ageron, Cyril Jaksic, Giulia Torriani, Erik Boehm, Ilona Kronig, Jilian A. Sacks, Margaretha de Vos, Frédérique Jacquerioz Bausch, François Chappuis, Laurent Kaiser, Manuel Schibler, Isabella Eckerle, for the Geneva Centre for Emerging Viral Diseases

**Affiliations:** Division of Infectious Disease, Geneva University Hospitals, Geneva, Switzerland; Geneva Centre for Emerging Viral Diseases, Geneva University Hospitals, Geneva, Switzerland; CRC & Division of Clinical-Epidemiology, Department of Health and Community Medicine, University of Geneva & University Hospitals of Geneva; Department of Microbiology and Molecular Medicine, University of Geneva, Geneva, Switzerland; Foundation for Innovative New Diagnostics, Geneva, Switzerland; Department of Primary Care, Geneva University Hospitals, Geneva, Switzerland; Laboratory of Virology, Division of Laboratory Medicine, Geneva University Hospitals, Geneva, Switzerland

## Abstract

**Background:** Antigen-detecting rapid diagnostic tests for SARS-CoV-2 offer new opportunities for the quick and laboratory-independent identification of infected individuals for control of the SARS-CoV-2 pandemic.

**Methods:** We performed a prospective, single-center, point of care validation of two antigen-detecting rapid diagnostic tests (Ag-RDT) in comparison to RT-PCR on nasopharyngeal swabs.

**Findings:** Between October 9^th^ and 23^rd^, 2020, 1064 participants were enrolled. The Panbio™Covid-19 Ag Rapid Test device (Abbott) was validated in 535 participants, with 106 positive Ag-RDT results out of 124 positive RT-PCR individuals, yielding a sensitivity of 85.5% (95% CI: 78.0–91.2). Specificity was 100.0% (95% CI: 99.1–100) in 411 RT-PCR negative individuals. The Standard Q Ag-RDT (SD Biosensor, Roche) was validated in 529 participants, with 170 positive Ag-RDT results out of 191 positive RT-PCR individuals, yielding a sensitivity of 89.0% (95%CI: 83.7–93.1). One false positive result was obtained in 338 RT-PCR negative individuals, yielding a specificity of 99.7% (95%CI: 98.4–100). For individuals presenting with fever 1-5 days post symptom onset, combined Ag-RDT sensitivity was above 95%.

**Interpretation:** We provide an independent validation of two widely available commercial Ag-RDTs, both meeting WHO criteria of ≥80% sensitivity and ≥97% specificity. Although less sensitive than RT-PCR, these assays could be beneficial due to their rapid results, ease of use, and independence from existing laboratory structures. Testing criteria focusing on patients with typical symptoms in their early symptomatic period onset could further increase diagnostic value.

**Funding:** Foundation of Innovative Diagnostics (FIND), Fondation privée des HUG, Pictet Charitable Foundation.

## Introduction

The 2019 novel coronavirus (SARS-CoV-2) which causes Coronavirus Disease 19 (COVID-19), has led to an unprecedented pandemic and a global public health crisis. Developing diagnostic strategies that are easily accessible, provide rapid results and are of low cost to allow use in resource limited settings is critical in order to control the pandemic. While RT-PCR based detection remains the standard for the detection of emerging respiratory viruses (1,2), the global need for virus detection methods has fueled research and development of diagnostic tests, including as Antigen-detecting rapid diagnostic tests (Ag-RDT), that can be performed at the point of care (POC) (3).

Such laboratory-independent tests could be key to detecting acutely infected and especially contagious individuals, allowing for control of the pandemic by quickly isolating individuals during their contagious period to prevent further transmission. These tests can also help overcome bottlenecks such as overwhelmed diagnostic laboratories and global shortages of PCR reagents, while being affordable (4). Ag-RDTs could enable mass screenings, which allow estimation of active infection prevalence, and confirming a COVID-19 diagnosis especially in settings where results are not available in a timely manner (5). Given that viral load, as measured by RNA levels, peaks near symptom onset (6–8) and contagiousness can begin days before symptom onset (9,10), it is expected that RDTs would have the highest sensitivity (SN) in those infected individuals that are most contagious. However, reported SNs of Ag-RDTs vary widely, and manufacturer reported SNs are often substantially higher than the SNs reported by independent assessments (11). The World Health Organisation (WHO) has published an Ag-RDT target product profile, aiming at SN ≥80% and a specificity (SP) of ≥97% (5,12). Thus, we sought to evaluate the performance of two commercially available Ag-RDTs through a prospective, single-center POC validation in comparison to RT-PCR for detecting SARS-CoV-2 using nasopharyngeal swabs (NPS) under real-life conditions.

## Methods

### Ethics

The study was approved by the Cantonal ethics committee (Nr. 2020-02323). All study participants and/or their legal guardians provided written informed consent.

### Setting

The study was performed in two geographically different testing centres in Geneva, run by our institution, the Geneva University Hospitals: one adjacent to the University hospital (“Sector E”) and the other located in another part of the city (“Sector G”) between October 9^th^ and October 23^rd^, 2020. Both centres are supervised by the same team, and did not differ in their infrastructure, so we analyzed them as a single centre study.

### Study design and participants

The primary objective of this prospective study was to assess the diagnostic accuracy (SN and SP) of the Ag-RDTs compared to the reference RT-PCR. All individuals presenting to the testing centres were informed about the study and enrolled if they consented. Participants were ≥16 years of age, with suspected SARS-CoV-2 infection according to the local governmental testing criteria. This included suggestive symptoms for COVID-19 and/or recent exposure to a SARS-CoV-2 positive person. Asymptomatic individuals were included if they were notified by the Swiss COVID-19 app about a contact offering the option to get tested on day 5 after contact, or if they received a notification from local health authorities (screening of people with high risk exposure in a cluster).

### Study procedures

For each participant, two NPS were collected. The first was a standard flocked swab placed in viral transport media (VTM), used routinely for viral genome detection by RT-PCR. The second NPS, provided in the Ag-RDT kit, was obtained from the contralateral nostril and was performed as recommended by the manufacturer. Both swabs were taken by the same trained nurse. All Ag-RDTs were performed immediately at the sample collection site. Adequate personal protective equipment was used while collecting the NPSs and performing the RDTs.

### Data collection

Clinical data were collected for each patient upon presentation with a questionnaire including the number of days post symptom onset (DPOS), known contact to a previous SARS-CoV-2 infected person, comorbidities and type of symptoms. The following symptoms were recorded: rhinorrhea (runny nose), odynophagia, myalgia, chills, dry cough, productive cough, red expectoration, fever (anamnestic), anosmia/ageusia (loss of smell or taste), gastrointestinal symptoms, asthenia, dyspnea, thoracic pain and headache.

Comorbidities included in the questionnaire were hypertension, cardiovascular disease, chronic pulmonary disease, diabetes, chronic renal failure, active cancer including lymphoproliferative disease, severe immunosuppression, immunosuppressive therapy, pregnancy, and obesity (BMI >40 kg/m^2^).

### Ag-RDT testing

The two validated Ag-RDTs were Panbio Covid-19 Ag Rapid Test device (Abbott Rapid Diagnostics) and Standard Q (SD Biosensor, Roche). Both Ag-RDTs were used as recommended by the manufacturers, using only materials provided by the manufacturers in the kits. Both assays were manually read, with two individuals reading the results separately after the indicated time. In case of discordant results, the two validators sought a consensus. All visible bands were considered a positive result. All Ag-RDT results were photographically documented.

### RT-PCR testing

All participants were tested by a dual target RT-PCR assay for SARS-CoV-2 (Cobas, Roche) using NPS in 3mL VTM. For further analysis, only cycle threshold (Ct) values for the E-gene were used. For calculation of viral loads (VL) as SARS-CoV-2 genome copy numbers per mL, a standard curve was obtained by using a quantified supernatant from a cell culture isolate of SARS-CoV-2. All VLs were calculated from the Ct-values, according to log10 SARS-CoV-2 RNA copies/ml = (Ct-44.5)/-3.3372 for Cobas (13,14).

### Statistics

We enrolled all patients who met the SARS-CoV-2 testing criteria, over a 2-week period. During the first and second week, 529 and 535 patients were enrolled respectively. The target sample size was 530, as it would have sufficient power to generate a 95% confidence interval (CI) with a lower bound above the WHO target of 80%, if the prevalence was 25% and the measured SP was ≥87.5%.

All continuous variables were presented by their mean ±standard deviation (SD) and median (interquartile range, IQR), categorical variables were presented by their frequencies and relative proportions. For comparisons of continuous variables, we used a nonparametric Mann-Whitney test due to small sizes; for comparisons of categorical variables, we either performed Chi^2^ or Fischer’s exact tests, depending on applicability.

To enable Ag-RDT result combination, we performed a Bayesian t-test on their sensitivities and specificities. To be able to conduct the t-test, the confidence intervals of both sensitivities and specificities were converted into standard deviation to allow for the t-test to be conducted. The test computes a Bayesian Factor (BF) that allows comparison of the probability of observing our data under H_0_ (both tests are equal in term of SN and SP) and H_1_ (both tests are different). All analyses were performed using STATA version intercooled 16 (Stata Corp., College Station, TX, USA). Statistical significance was defined as p<0.05 (two-sided).

## Results

Between October 9^th^ and October 23^rd^, 2020, 1064 participants were enrolled and included in the analysis. 535 participants were tested with the Panbio Ag-RDT from October 09^th^ to 16^th^ and 529 participants were tested with the Standard Q Ag-RDT from October 19^th^ to 23^rd^, 2020. The demographic and clinical characteristics of the study population are shown in **Table 1**. The mean age of the study participants was 34.9 years (SD ±10.9) with 53.8% being female. The mean DPOS to testing was 2.7 (SD ±1day). Overall, 29.6% of participants were positive by RT-PCR with a mean Ct-value of 22.5 (SD ±5.1), corresponding to a VL of 1.8E7 SARS-CoV-2 copies/mL. Most patients (97.8%) were symptomatic upon presentation at the testing centre, with only 3 reporting no symptoms. Symptoms information was missing for 4 patients. The study population tested with the Standard Q vs. was younger than that tested with the Panbio assay (34.9 ±10.9 vs 38.5 ±13.6 years, respectively, p<0.001) and DPOS differed slightly (2.9±1.5 vs. 2.6±2.0 days, respectively, p=0.0125). Ct-values did not differ significantly between the two groups (p=0.450): 22.6 (SD ±4.9) in the Standard Q group vs. 22.4 (SD ±5.4) in the Panbio group, corresponding to 1.7E7 and 1.9E7 SARS-CoV-2 RNA copies/mL, respectively. The RT-PCR positivity rate was 23.2% and 36.1% for the population tested with the Panbio and the Standard Q, respectively, corresponding to an increase in the overall PCR positivity rate and reflecting the rapidly increasing local incidence during the time of this study.

**Table 1.**
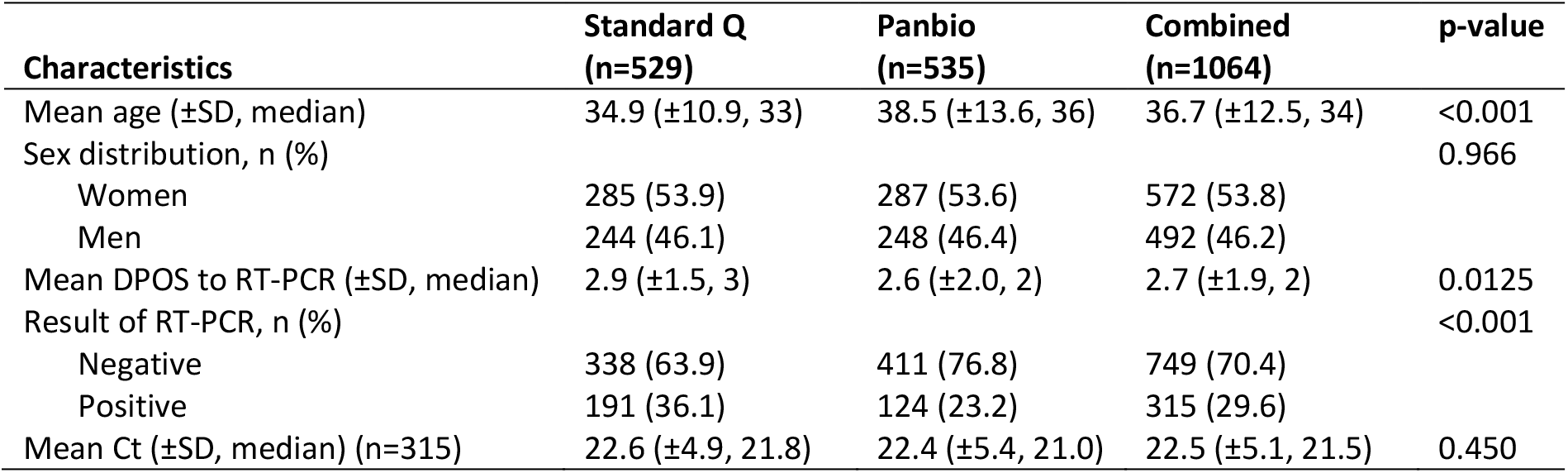
Characteristics of the study population. SD, standard deviation; RT-PCR, reverse transcription polymerase chain reaction; DPOS, days post symptom onset; Ct, cycle threshold

| Characteristics | Standard Q<br>(n=529) | Panbio<br>(n=535) | Combined<br>(n=1064) | p-value |
| --- | --- | --- | --- | --- |
| Mean age (±SD, median) | 34.9 (±10.9, 33) | 38.5 (±13.6, 36) | 36.7 (±12.5, 34) | <0.001 |
| Sex distribution, n (%) |  |  |  | 0.966 |
| Women | 285 (53.9) | 287 (53.6) | 572 (53.8) |  |
| Men | 244 (46.1) | 248 (46.4) | 492 (46.2) |  |
| Mean DPOS to RT-PCR (±SD, median) | 2.9 (±1.5, 3) | 2.6 (±2.0, 2) | 2.7 (±1.9, 2) | 0.0125 |
| Result of RT-PCR, n (%) |  |  |  | <0.001 |
| Negative | 338 (63.9) | 411 (76.8) | 749 (70.4) |  |
| Positive | 191 (36.1) | 124 (23.2) | 315 (29.6) |  |
| Mean Ct (±SD, median) (n=315) | 22.6 (±4.9, 21.8) | 22.4 (±5.4, 21.0) | 22.5 (±5.1, 21.5) | 0.450 |

The overall test performance for the Standard Q was 89.0% SN (95%CI: 83.7-93.1) and 99.7% SP (95%CI: 98.4-100%). The overall test performance for the Panbio assay was 85.5% SN (95%CI: 78.0-91.2) and 100% SP (95%CI: 99.1-100) (**Table 2**).

**Table 2.** Overall SN, SP, positive and negative predictive value of Standard Q and Panbio SARS-CoV-2 Ag-RDT. Positivity rate at the time of study for Standard Q was 36.1% and at the time of study for Panbio was 23.2%.

| Characteristics | Standard Q | Panbio | Combined |
| --- | --- | --- | --- |
| SN, % (95%CI) | 89.0 (83.7-93.1) | 85.5 (78.0-91.2) | 87.6 (83.5-91.0) |
| SP, % (95%CI) | 99.7 (98.4-100) | 100 (99.1-100) | 99.9 (99.3-100) |
| Positive predictive value, % (95%CI) | 99.4 (96.8-100) | 100 (96.6-100) | 99.6 (98.0-100) |
| Negative predictive value, % (95%CI) | 94.1 (91.2-96.3) | 95.8 (93.4-97.5) | 95 (93.3-96.5) |

Ct-values of samples with positive Ag-RDT results ranged from 14.2-34.0 and 14.4-34.2 for Panbio and Standard Q (p=0.1766), respectively, while Ct-values of samples of samples tha tested falsely negative by Ag-RDT ranged from 16.0-39.7 and 19.8-37.4 (p=0.7998), respectively. Median Ct-values of Ag-RDT positive samples (Panbio: 20.4, IQR: 18.1-23.8; Standard Q: 21.2, IQR 18.6-24) were lower than those of Ag-RDT negative samples (Panbio: 30.5, IQR: 27-35.9; Standard Q: 30.4, IQR: 25.7-33.9) (**Figure 1**). Furthermore, we evaluated overall Ag-RDT results in relation to Ct-values/viral load as well DPOS (**Figure 2**). False-negative results occurred in both assay across all DPOS.

**Figure 1.**
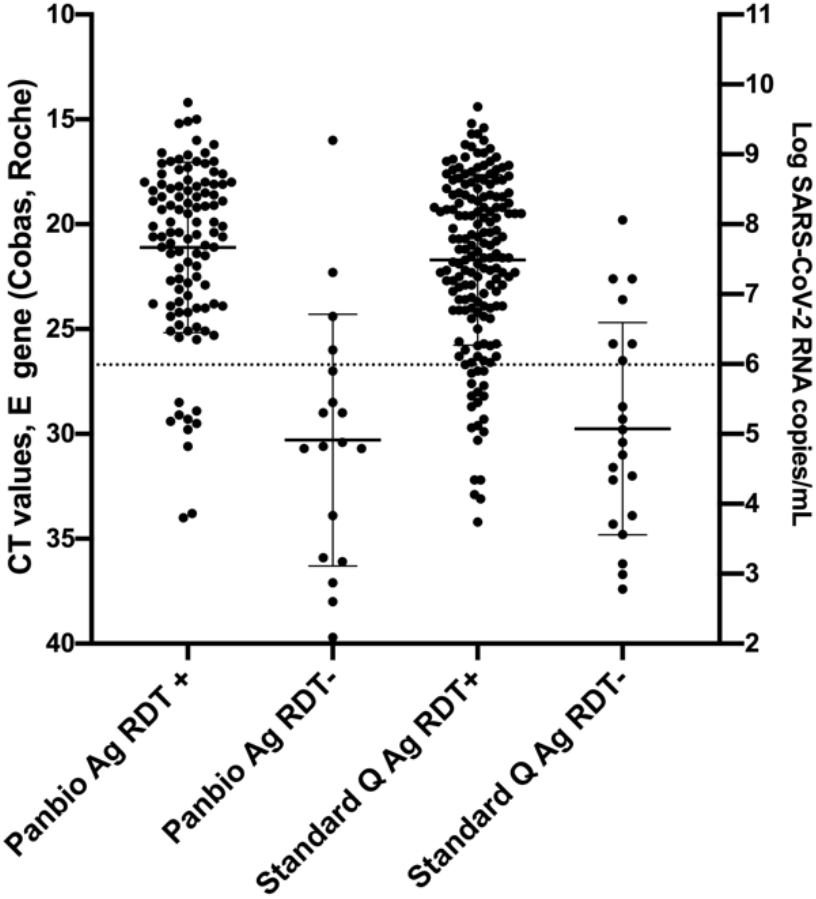
Ct values, viral load and Ag-RDT results for RT-PCR-positive individuals tested with Standard Q (n= 191) and Panbio (n=124). Horizontal bars represent median and standard deviation. Dotted line: Ct value of 26.7 or 1E6 SARS-CoV-2 RNA copy numbers/mL.

**Figure 2.**
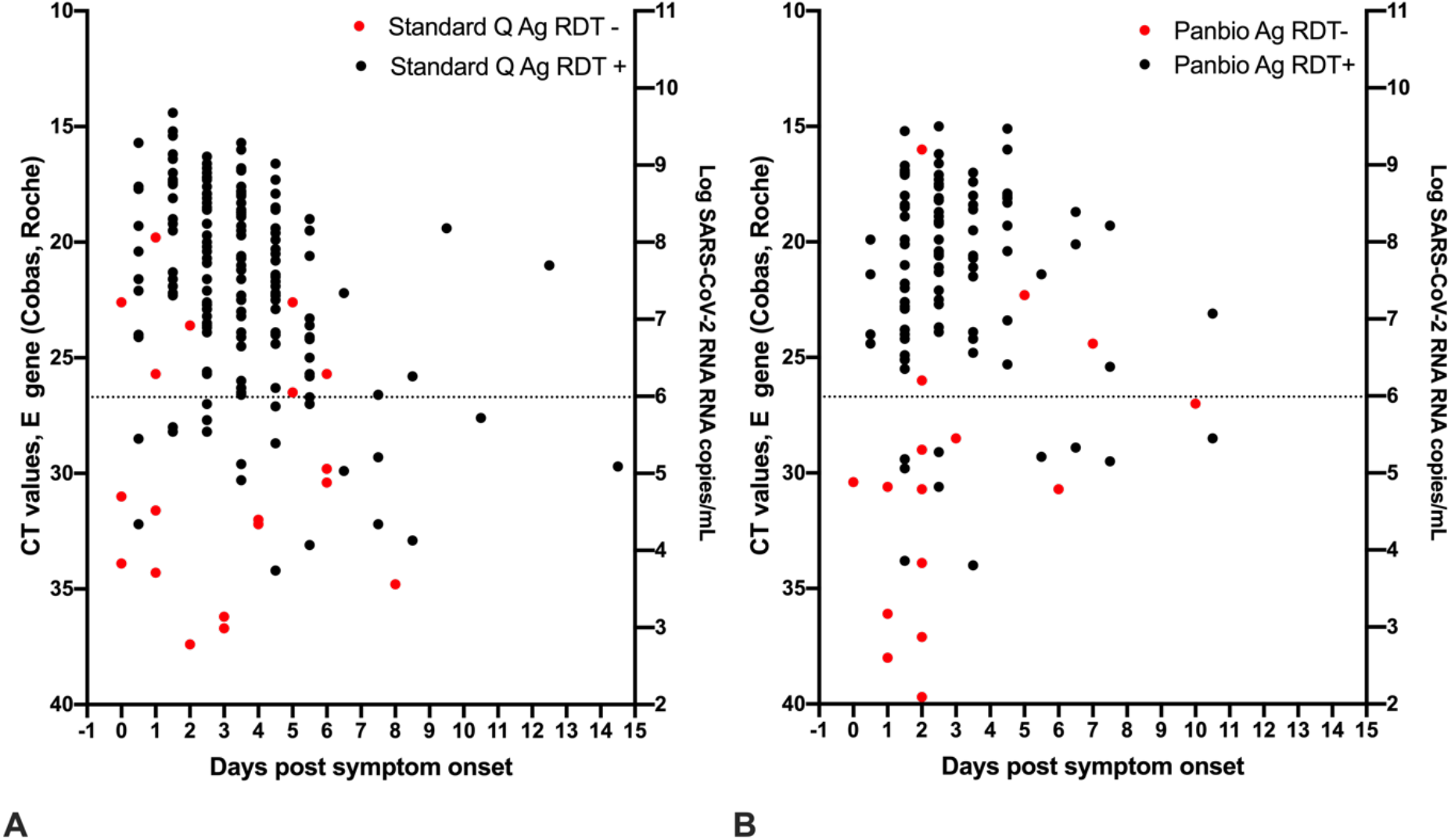
Ct values, viral load, days post symptom onset and Ag-RDT results for 190 patients tested with Standard Q (**A**) and 116 patients tested with Panbio (**B**) for which information on day of symptom onset was available. Dotted line: Ct value of 26.7 or 1E6 SARS-CoV-2 RNA copy numbers/mL.

We compared SN and SP between the two Ag-RDTs and concluded that we could accept, with high probabilities, (respectively likelihood ratio of BF_01_=10.2 and 11.9) the hypothesis of equivalent SN and SP. Based on this, a combined SN of 87.6% (95%CI: 83.5-91.0) and a combined SP of 99.9% (95%CI: 99.3-100) for both Ag-RDTs were calculated with a positive predictive value of 99.6% (95%CI: 98.0-100) and a negative predictive value of 95.0% (95%CI: 93.3-96.5). In order to identify subpopulations in which maximal SN could be reached with these tests, we analyzed SN by DPOS, Ct-values as determined by RT-PCR, type of symptoms, comorbidities, and previous contact with a confirmed SARS-CoV-2 infection.

Combined SN varied according to Ct-values: it was highest in samples with low Ct-values, with a SN of 98.4% (95% CI: 94.2-99.8) for Ct ≤20 (≤1.0E8 SARS-CoV-2 copies/mL), decreased slightly to 95.5% (95%CI: 89.9-98.5) for 20 < Ct ≤25 (<1.0E8 SARS-CoV-2 copies/mL ≤ 3.2E6), dropped further to 89.9% (95%CI: 86.0-93.0) for Ct ≤35 (≤3.2 E3 SARS-CoV-2 copies/mL) and was lowest (only 40.9% (95%CI: 20.7-63.6)) for 30< Ct ≤35 (<1.0E5 SARS-CoV-2 copies/mL ≤3E3) (**Figure 3**). The SN for all samples with a Ct value ≤26.7 (≤1E6 SARS-CoV-2 copies/mL), an assumed cut-off for presence of infectious virus, was 95.7% (95%CI: 92.4-97.8).

**Figure 3.**
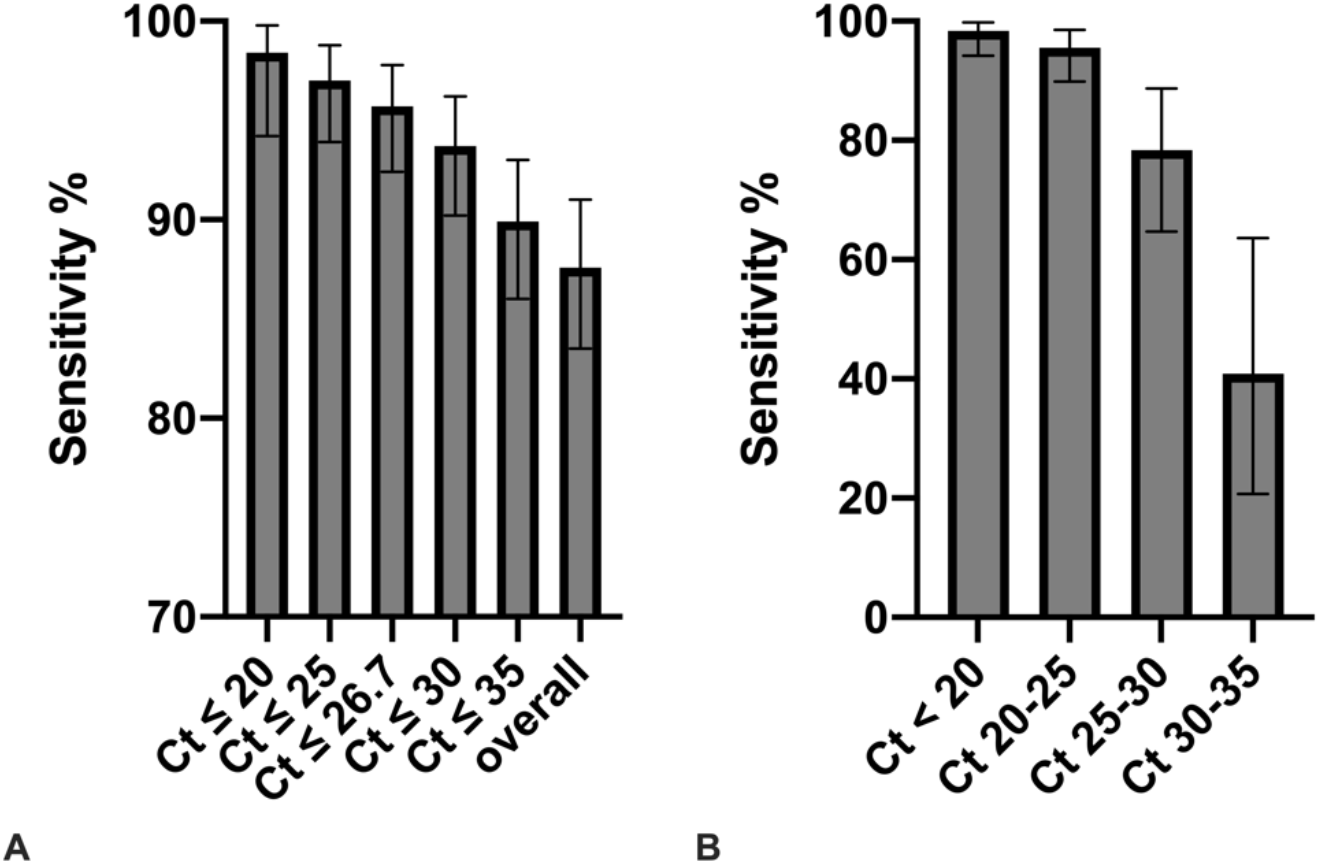
**A**. Combined SN of the two Ag-RDTs according to Ct-values of the RT-PCR. **B**. Combined SN of the two Ag-RDTs according to subgroups of Ct-values of the RT-PCR. Ct values correspond to the following SARS-CoV-2 RNA copy numbers/mL: Ct 20: 1.0E8; Ct 25: 3.2E6; Ct 26.7: 1E6; Ct 30: 1.0E5, Ct 35: 3.2E3.

SN increased with DPOS, from 88.2% at 0 DPOS (95%CI: 63.6-98.5) to 94.3% (95%CI: 84.3-98.8, p=0.030) at 1 DPOS, and remained high until 5 DPOS. The highest SN was seen between 1 DPOS and 4-5 DPOS, ranging from 94.3% (95%CI: 84.3-98.8) to 94.8% (95%CI: 85.6-98.9), with a decline after 5 DPOS (**Figure 4A**).

**Figure 4.**
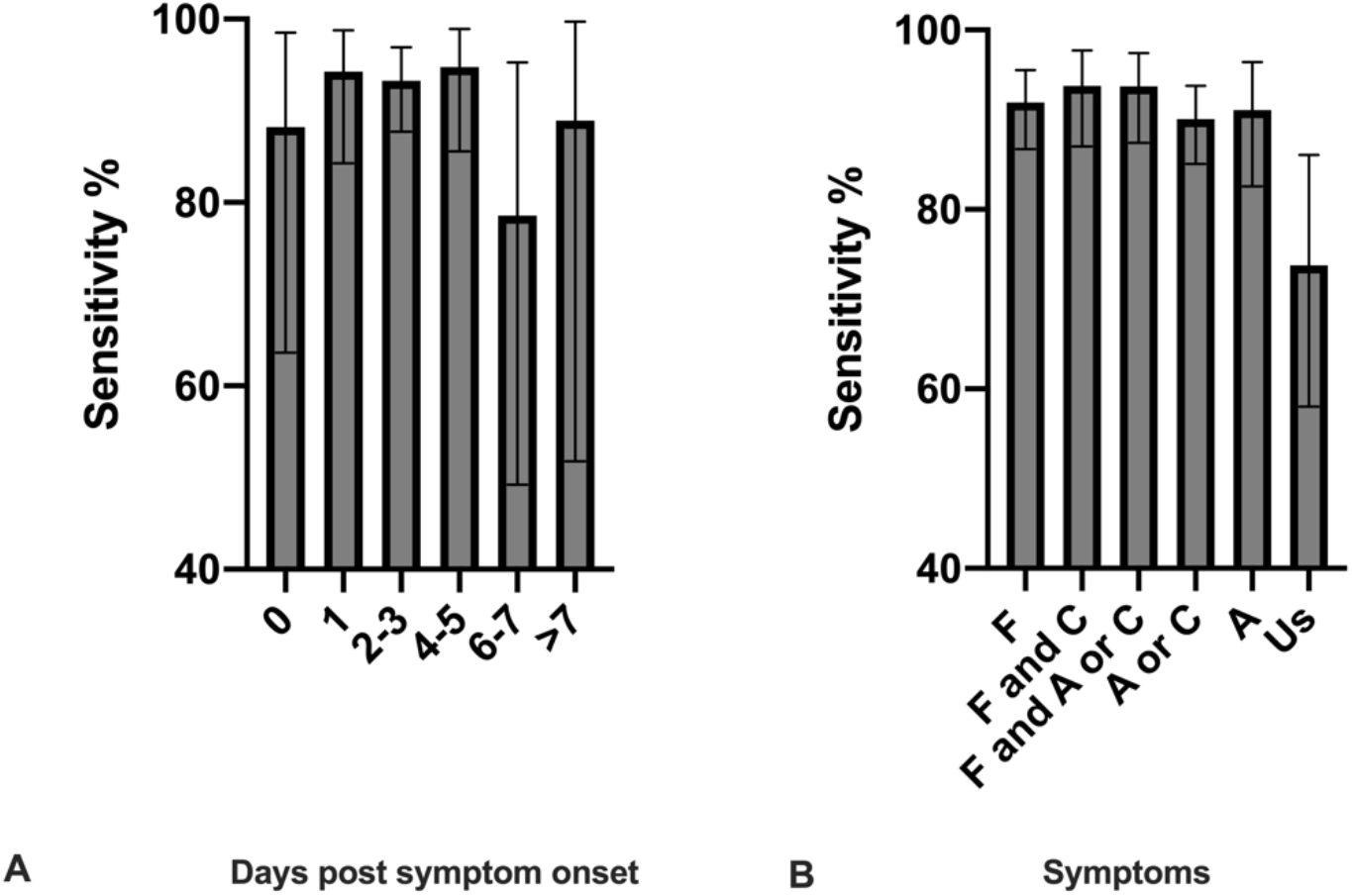
**A**. Combined SN of the two Ag-RDTs according to days post symptom onset. Number of patients per category: Day 0, n= 17; day 1, n=53, day 2-3, n=135; day 4-5, n=58; day 6-7, n=14; > 7 days, n= 9). **B**. Combined SN of the two Ag-RDTs according to symptoms F, fever/chills; C, cough, A, anosmia/ageusia (loss of smell or taste), Us, unspecific symptoms (all other symptoms excluding fever/chills, cough and anosmia/angeusia). Number of patients per category: F, n= 172; F and C, n= 97; F and A or C, n=111; A or C, n= 202; A, n= 79; Us, n=42.

Additionally, we analyzed SN according to specific symptoms, differentiating between typical COVID-19 symptoms (fever/chills, cough and anosmia/ageusia) and more non-specific symptoms of respiratory infection (all other symptoms reported). The highest SN of 93.8% (87.0-97.7) was observed for patients presenting with fever/chills and cough at the time of testing, followed by patients presenting with anosmia/ageusia or cough and fever/chills with a SN of 93.7% (95%CI: 87.4-97.4), but only 73.8% (95%CI: 58.0-86.1) in patients presenting with non-specific signs (**Figure 4B**). No difference in SN was seen between patients with (89.3%, 95%CI: 71.8-97.7, n=28) or without (87.5%, 95%CI: 83.1-91.1, n=287) comorbidities (p=0.999). Typical symptoms were more frequent in patients with comorbidities (100%, 15/15 patients) than in patients without comorbidities (86.5%, 96/138 patients) (p=0.012), however sample size was small. No difference was seen in patients with or without contact with a recently positive case (p=0.065). We further analysed by DPOS, and found that the highest SN was seen in patients with fever/chills and presenting between 1 and 5 DPOS, at 95.7% SN (95%CI: 91.0-98.4).

## Discussion

This study provides an independent, POC validation of two commercial Ag-RDTs in comparison to SARS-CoV-2 RT-PCR and according to demographic and clinical information. This combined validation of two similar assays provides performance data in a real-life high incidence test setting with an approach aimed at an immediate implementation solution.

Both RDTs performed well with an overall SN of 87.6% (95%CI: 83.5-91) and a very high SP of 99.9% (95%CI: 99.3-100) in our test setting during a time of very high SARS-CoV-2 weekly incidence (375/100,000 to 824/100,000 inhabitants) and a SARS-CoV-2 RT-PCR positivity rate >20%. SN was higher in sub-populations with earlier DPOS numbers and characteristic COVID-19 symptoms. Importantly we highlight that on day 0 of symptoms, the SN may be lower than during the subsequent days, and as expected, the sensitivity drops rapidly when the Ct-values increase above a threshold of 30, mostly after 6-7 days. These results suggest that with increasing availability, Ag-RDTs can enable rapid and reliable identification of SARS-CoV-2 cases and hold a promise for more efficient control of the current pandemic, independent of existing diagnostic structures.

The highest VL and thus the highest SARS-CoV-2 transmission probability occurs within the first week of symptom onset, with VLs peaking shortly before or at the time of symptom onset (6). Culturable virus has been predominantly found in the first week after symptom onset, up to a VL in the range of 1E6 copies/ml (14–17). This cut-off was also chosen by WHO in their Ag-RDT target product profile guide for SARS-CoV-2 diagnostics (12). The SN of the Ag-RDTs validated here, for patients presenting with a VL compatible with contagiousness, was 95.7%. Correspondingly, the highest Ag-RDT SN was also observed at early DPOS numbers and in patients with low Ct-values, again suggesting reliable identification of contagious individuals. Our findings at the POC are in line with other validations performed across countries and at different SARS-CoV-2 prevalence, although study designs and specimens used varied considerably between studies. Standard Q was reported to have SNs between 70.6-88.7%, while SP remained high throughout these studies between 97.6-100% (18–24). A clinical study performed similarly to ours in a much lower-incidence setting (<1% RT-PCR positivity rate), found a SN/SP of 76.6%/100%, using a mixture of NPS and combined oro- and naso-pharyngeal swabs from a total of 2417 participants with 47 RT-PCR positive samples yielding 36 Ag-RDT (18).

For Panbio, other studies have reported SNs ranging from 73.3-91.7% with SP in the range of 94.9-100% (25–28). Notably, the highest reported SN of 91.7%/98.9% comes from a study using 1,406 frozen archived NPS specimens of which 951 were positive by RT-PCR. However here tests were not done at the POC, and although the use of frozen samples is possible for RDTs (depending on manufacturers recommendation and on viral transport medium used), it is not their intended use (19,25). It is also unknown if a freeze-thaw cycle can affect the accessibility of viral antigens for Ag-RDT purposes.

While significant variation in overall SN is observed for the validated Ag-RDTs across studies, there is remarkable similarity when comparing samples with Ct-values in the same range – although caution must be exercised equating Ct-values to VLs across different studies. When considering Ct-values of <25, the Standard Q test was reported to have a SN of 100% (18), while the Panbio was reported to have a SN of 97.1% (27) or 98.2% (25), which is in agreement with our results of 97%. In contrast to most other validations, we did observe some cases of false-negative Ag-RDT results in patients with low Ct-values/high VLs across a range of DPOS. It is likely that these patients are contagious and could transmit SARS-CoV-2, with VLs that are compatible with culturable virus.

We did not find any validation of Ag-RDTs that has analyzed SN based on type of symptoms, which could be an additional factor for testing algorithms. Our data suggests that the best SN is found in symptomatic individuals with symptoms suggestive of COVID-19, early in the disease between day 1 and 5. The presence of only non-specific symptoms of respiratory illness corresponded to lower SN, however, few study participants had no COVID-19 specific symptoms at all.

Our study has several strengths. The Ag-RDTs were performed at the POC in parallel to RT-PCR, and is one of the largest in terms of RT-PCR positive individuals. The test population was mainly young adults without comorbidities, who mostly had typical COVID-19 symptoms that were still mild enough not to require hospitalization. This represents a population screened for public health intervention, and not for diagnostic purposes in a hospital setting; and the majority of SARS-CoV-2 infected individuals at the current phase of the epidemic in the community, which is an important group for early identification to limit viral transmission. Thus, the results of our study support implementing the Ag-RDTs in a decentralized manner for community testing, which could significantly alleviate the burden on diagnostic laboratories and hospital staff.

Although Ag-RDTs are less sensitive than RT-PCR and, in our study, false-negative Ag-RDT results were also seen in patients with high VL, the public health benefit of quickly identifying a large proportion of SARS-CoV-2 positive individuals in the community would still outweigh the disadvantages of occasional missed diagnoses (29). Repeatedly testing and following general recommendations like self-isolation for symptomatic individuals, even after a negative Ag-RDT would probably largely prevent further spread from individuals who had false-negative results. In a high prevalence situation like ours, positive Ag-RDTs have a high positive-predictive value, while negative results are less reliable, and thus negative-results should not be used for rule-out purposes or for any kind of reduced infection prevention measures. The lower SN found at 0 DPOS was probably due to a still increasing VL. Re-testing with an Ag-RDT or RT-PCR, in case of persisting symptoms, could overcome this limitation.

Furthermore, our validation showed very high SP, with only one false-positive Ag-RDT result in an overall sample of 315 RT-PCR positive patients. Interestingly, the patient with this putative false-positive result returned 3 days later because of persisting respiratory symptoms, and then tested positive for SARS-CoV-2 by RT-PCR.

During the time this study was conducted, the Canton of Geneva was experiencing a severe second wave of SARS-CoV-2 infections, with a very high incidence as well as high RT-PCR positivity rates, thus extrapolating the findings of our study to low prevalence settings must be done with caution. Furthermore, our study population consisted mainly of young symptomatic individuals in an outpatient setting, thus diagnostic performance in other groups needs further validation. All diagnostic testing was performed on NPS collected by trained nurses, thus the use of the test with other materials (e.g. anterior nares swabs) or in other settings (e.g. self-testing) needs to be further validated, however a first study on anterior nasal samples showed promising results (30). Due to logistical and ethical reasons, we were not able to perform paired comparisons of both Ag-RDTs at the same time by taking three separate NPS, which would have helped to control for prevalence variation and inter-cohort variability. Nonetheless, we used robust methods to confirm that the SN and SP of both Ag-RDTs was equivalent, and further combined results of both Ag-RDTs.

In conclusion, we show good diagnostic accuracy of both Ag-RDTs, especially for rule-in purposes of infected individuals and in testing patients with defined clinical criteria. The SN for identification of SARS-CoV-2 infected individuals during the period of highest infectiousness, the rapidity of results as well as the independence from existing laboratory structures make these Ag-RDTs promising tools for SARS-CoV-2 infection control in the community.

## Data Availability

Original data are available upon request according to the regulations of the ethical approval.

## Acknowledgements

We thank all nurses and staff at the testing Centre Sectors of our institution as well as the patients for their willingness to participate in the study. We thank Catia Machado-Delgado for excellent technical assistance and Stéphanie Baggio for help with data analysis.

